# Endemic transmission and international introduction of Monkeypox virus in Southern Brazil between 2022-2023

**DOI:** 10.1101/2024.06.22.24309232

**Authors:** Fernanda Marques Godinho, Thales de Lima Bermann, Mayara Mota de Oliveira, Regina Bones Barcellos, Amanda Pellenz Ruivo, Viviane Horn de Melo, Franciellen Machado dos Santos, Milena Bauermann, Taina Machado Selayaran, Taina dos Santos Soares, Patrícia Sesterheim, Ludmila Fiorenzano Baethgen, Fernanda Maria Da Rocha, Karine Medeiros Amaral, Fernanda Crestina Leitenski Delela, Renata Petzhold Mondini, Sabrina Vizeu, Tatiana Schäffer Gregianini, Ana Beatriz Gorini da Veiga, Gabriel da Luz Wallau, Richard Steiner Salvato

## Abstract

Mpox is a zoonotic viral disease caused by the monkeypox virus (MPXV). Human cases have been mainly restricted to the African continent until the worldwide multi-country outbreak unfolded in 2022. We reconstructed epidemiological links of MPXV infections using genomic epidemiology in Rio Grande do Sul State, southern Brazil, during 2022 and 2023. We detected five well-supported clades, three representing local transmission chains that were mostly restricted to the 2022 virus spread, one supported year-long maintenance encompassing samples from 2022 and 2023, and one new importation from Europe in 2023. Our results provide new insights into the geographic extent of community transmission and its association with viral diversity during the more pronounced 2022 mpox upsurge and during the following lower incidence endemic transmission phase. These findings highlight the power of continued genomic surveillance to uncover hidden transmission chains to understand viral dynamics and inform public health responses. The detection of sustained endemic transmission in the state is important to guide targeted control measures to curtail further community and international transmission and highlight the need for strengthening genomic surveillance.

## Introduction

Mpox (formerly named monkeypox) is a zoonotic disease caused by the monkeypox virus (MPXV), an enveloped double-stranded DNA virus belonging to the *Orthopoxvirus* genus of the *Poxviridae* family (1). The first reported human mpox case was detected in 1970 in the Democratic Republic of the Congo, and since then endemic transmission has been reported in the African continent (2). Before 2022, several mpox human cases in non-African countries were registered. However, these events were generally linked to patient travel history to endemic areas, and limited onward human-to-human transmission was registered outside of the African continent (3,4). In July 2022, following the notification of a series of mpox cases primarily across Europe and the subsequent widespread transmission to multiple countries, the World Health Organization (WHO) declared a Public Health Emergency of International Concern (PHEIC) (5–7). The outbreak was marked by a rising number of reported cases across 117 countries, and at the end of 2023, it surpassed 97,000 confirmed cases worldwide (https://worldhealthorg.shinyapps.io/mpx_global/).

According to the recently introduced nomenclature, the MPXV genomes identified during the 2022 outbreak belong to the IIb clade (formerly named ‘West African’ clade). Strains belonging to this clade are known to typically cause milder symptoms and a lower mortality rate compared to infection by strains of clade I, previously named ‘Congo Basin’ clade (8–10). Clade IIb demonstrated sustained human-to-human transmission, which led to the designation of a new subclade named ‘hMPXV1’ within Clade IIb. A lineage scheme was adopted in this subclade (starting with ‘A’ and subsequent clades are designated as ‘A.1’, ‘A.2’, and ‘A.1.1’), similar to the SARS-CoV-2 lineage system. Most of the sequences from the mpox 2022 worldwide outbreak belong to the B.1 lineage (11).

Changes in the MPXV genome could impact the virus fitness by influencing factors such as transmissibility and virulence (12), and these mutations can also impact diagnostic detection (13). Consequently, genomic surveillance emerges as a crucial tool in supporting public health strategies aimed at understanding the transmission dynamics and mitigating virus transmission. This approach helps in defining and prioritizing public health interventions in high-risk areas/populations by tracking the geographic spread, timing, and evolution of virus lineages (14). Additionally, it aids in the identification of emerging variants that may exhibit altered virulence or transmissibility. Furthermore, the utility of genomic data extends to optimizing diagnostic tests and guiding vaccine development efforts. Understanding the genetic composition of circulating viruses through genomic surveillance contributes significantly to risk assessment and outbreak control strategies. The ability to make accurate inferences regarding outbreak trajectories enables the implementation of tailored control measures, thus enhancing overall preparedness and response capabilities (15).

During the 2022 mpox outbreak, Brazil ranked among the countries most affected by the disease worldwide, with 95,226 confirmed cases reported by March of 2024 - the period of the last reported data (https://worldhealthorg.shinyapps.io/mpx_global/). Rio Grande do Sul (RS), the southernmost state of Brazil, accounted for 3% of all cases country-wide, with a total of 337 confirmed cases as of late 2023 (16). In this study, we reconstructed the epidemiological link of cases in Rio Grande do Sul through the genomic characterization of mpox virus shedding new light on long-term local viral transmission and new international introduction in Brazil and the state.

## Methods

### Patient samples and MPXV detection

Mpox is a notifiable disease in Brazil, meaning that healthcare authorities must be informed of any suspected or detected cases. Suspected cases are defined as individuals of any age who experience a sudden onset of acute rash suggestive of mpox (typically deep and well-defined, often with central umbilication, and progress through specific sequential stages – macules, papules, vesicles, pustules, and crusts), whether singular or multiple, occurring anywhere on the body (including the genital region), with or without associated lymphadenopathy or reported fever.

As part of the Rio Grande do Sul State Health Department mpox diagnosis routine, our laboratory received samples collected from suspected individuals across the state. Viral DNA was extracted from clinical specimens (swabs of lesion surface, exudate, or lesion crusts) with the automatic extractor *Loccus Extracta 96* (São Paulo, Brazil) using the *Extracta Kit Fast–DNA and RNA Viral*, according to the manufacturer’s instructions. Mpox diagnosis was performed based on a Real-Time PCR assay for the specific detection of MPXV West African clade according to the protocol previously published (17) on the *Bio-Rad CFX Opus 96* (CA, USA) instrument. Confirmed cases were identified based on an amplification Cycle Threshold (Ct) of up to 38.

### Genome sequencing and assembly

Among 337 confirmed cases, a subset of 53 clinical specimens, sampled between July 2022 and November 2023 and exhibiting a Ct value of less than 29, were randomly selected for whole-genome sequencing to characterize their genomic composition. The sequencing was performed using an amplicon-based protocol described elsewhere (18), consisting of generating tailed amplicons of MPXV genome that went through library preparation using Illumina kits (*Illumina DNAPrep or Illumina COVIDSeq*). The libraries were then sequenced on the *Illumina MiSeq* platform using *Illumina MiSeq V3 600 cycle* kits. Raw sequencing data in FASTQ format were processed through the *ViralFlow* pipeline (https://viralflow.github.io/), providing a comprehensive workflow for referenced-based assembly, quality control, variant calling, and consensus sequence generation (19).

### Phylogenetic analysis

Consensus sequences were retrieved and used to reconstruct phylogenetic trees using a maximum likelihood approach implemented on *IQTREE2* (20), clade/lineage assignment, and mutation profiling on *NextClade v2.14.1* (https://clades.nextstrain.org/) (21). For phylogenetic analysis, the 53 sequenced samples were analyzed within the context of a larger dataset, comprising 848 MPXV sequences worldwide, subsampled using *NextClade* along with 216 high-quality MPXV sequences from Brazil obtained from *GISAID* (**Supplementary Table 1**). ModelFinder was employed to identify the most suitable nucleotide substitution model (22). The robustness of the tree was assessed with both 1000 fast-bootstrap replicates and Shimodaira-Hasegawa approximate likelihood ratio test (SH-aLRT) support, the final tree was visualized in FigTree (https://tree.bio.ed.ac.uk/software/figtree/).

## Results

Rio Grande do Sul State reported its first mpox case in May 2022. By the end of that year, there were 328 confirmed cases out of 2,529 suspected cases. Analysis of the distribution of cases by notification date demonstrated that the peak of infections occurred in August and September, representing over 67% of the total cases of the year (Figure 1). By contrast, only nine cases were confirmed in 2023 out of 407 suspected cases. The epidemiological profile of confirmed and sequenced mpox cases in this study revealed a male predominance (n=300, 89.02%), with a median age of 34 years (range 0-73 years). Of these, 62.91% (n=212) were identified as cis men and 52.82% (n=178) were homosexual. With regards to self-reported ethnicity, 69.70% (n=230) were white (**Table 1**).

**Figure 1.**
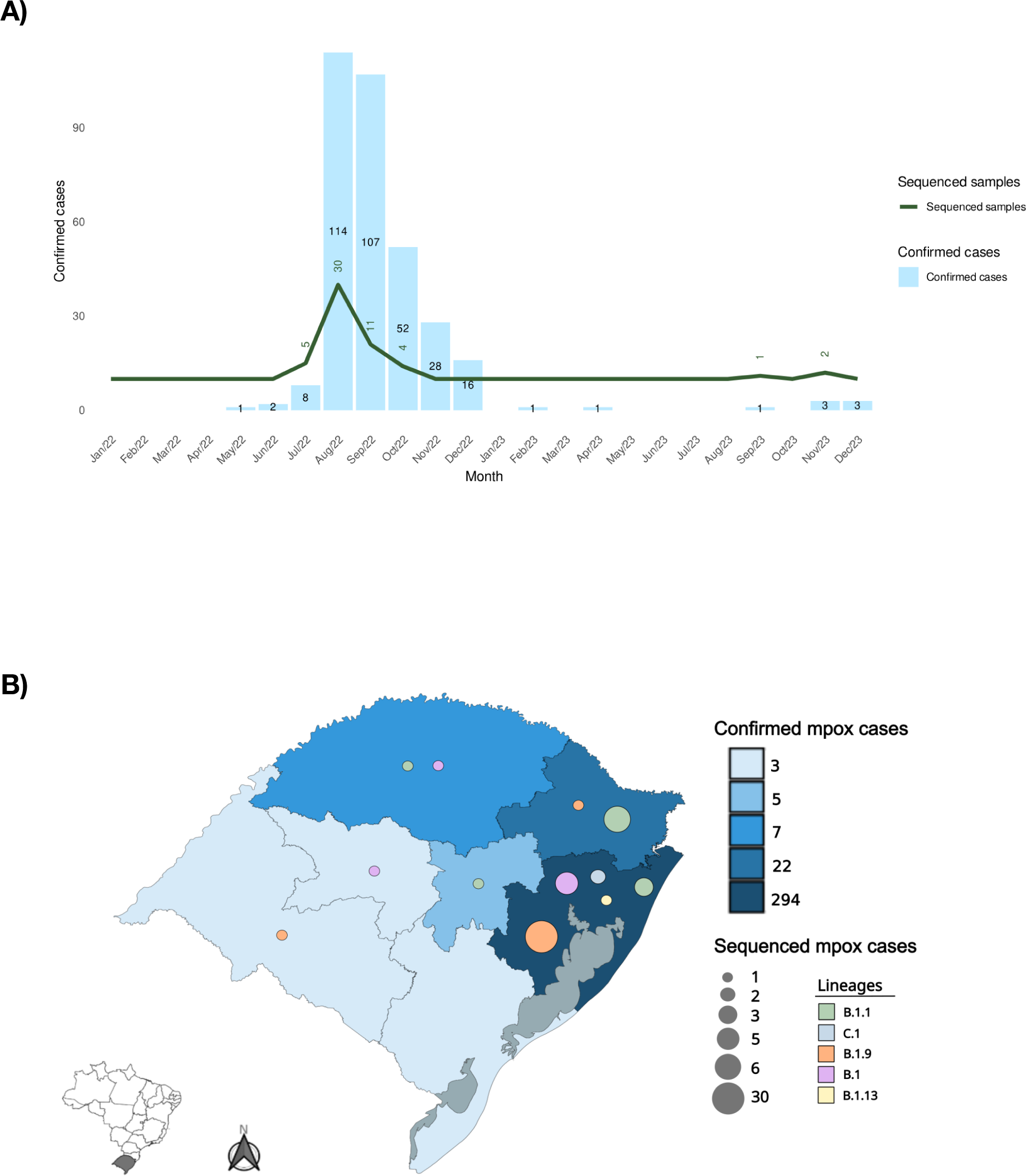
**A)** Distribution of mpox Cases in Rio Grande do Sul State, Brazil (2022–2023) and number of sequenced samples by month of notification. **B)** Map showing the number of confirmed mpox cases and mpox-sequenced cases among the different Rio Grande do Sul State regions.

**Table 1.**
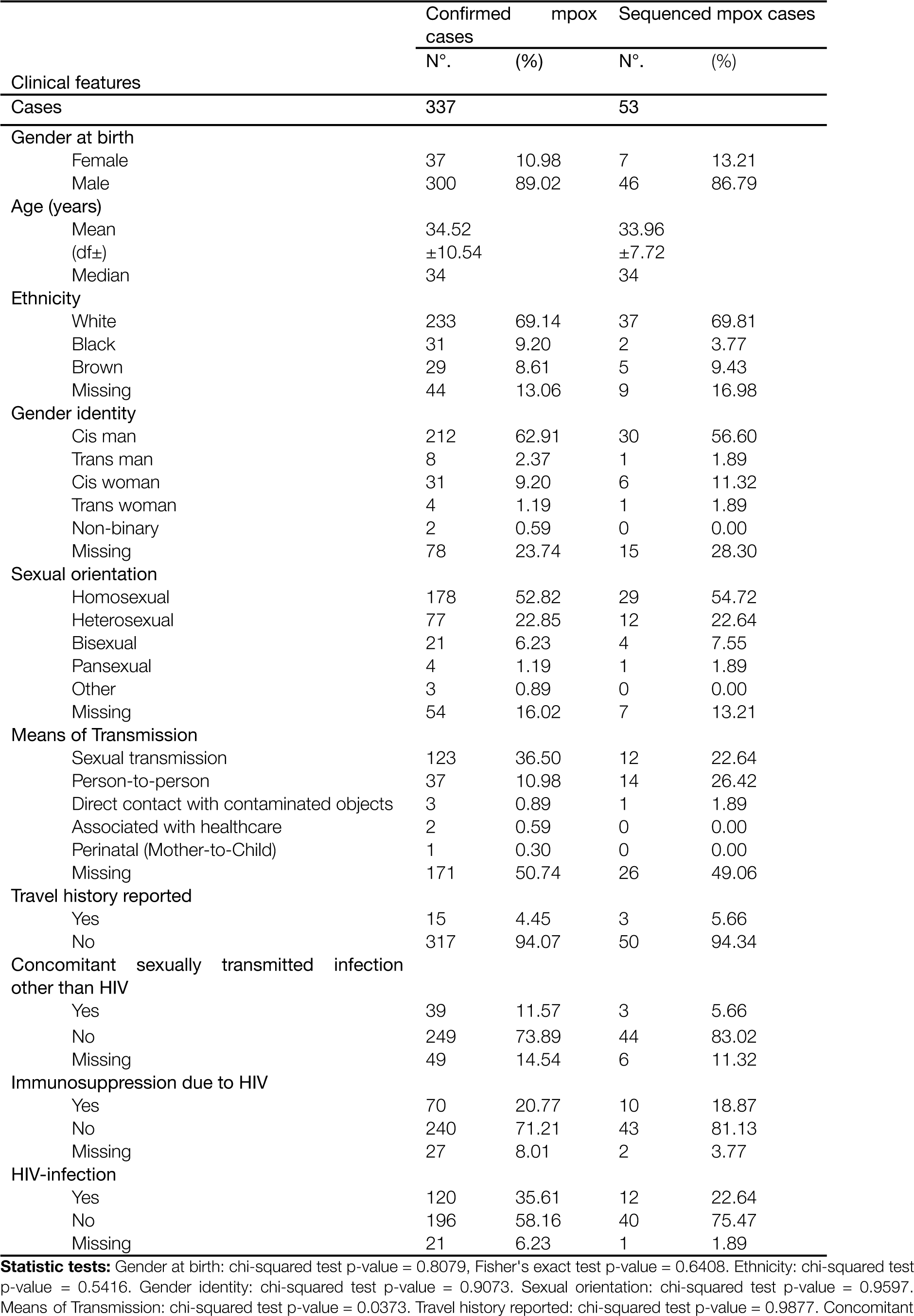
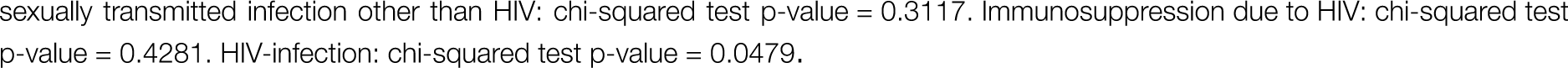
Clinical and epidemiological characteristics of confirmed mpox cases and sequenced cases of mpox in Rio Grande do Sul, Brazil, 2022 and 2023.

Among the confirmed mpox cases, the most frequent symptom was skin rash (72.70%), followed by fever (54.90%), and genital/perineal lesions (41.25%). Other common clinical features included headache (31.16%), myalgia (27.30%), chills (25.82%), asthenia (24.93%), sore throat (23.74%) and adenomegaly (23.15%). Less frequently reported symptoms included oral lesions (11.57%), back pain (11.57%), nausea (13.39%), penile edema (10.39%), and proctitis (9.79%). Most mpox cases reported in Rio Grande do Sul did not require hospitalization (93.64%), and no deaths were recorded in the state. The 21 (6.36%) patients who required hospitalization were primarily hospitalized for isolation or clinical management. Of these, two pregnant women were hospitalized for monitoring purposes and only one patient necessitated intensive care. All hospitalized patients evolved to cure and were discharged.

Among confirmed cases, sexual transmission was the most commonly reported probable route (n=123, 36.50% overall; n=12, 22.64% sequenced), followed by non-sexual person-to-person transmission (n=37, 10.98% overall; n=14, 26.42% sequenced). Person-to-person transmission was considered the probable route of infection when the person seeking care had close, non-sexual physical contact with symptomatic cases within 21 days prior to the onset of signs and symptoms.

A substantial proportion of cases had an unknown probable transmission route (n=171, 50.74% overall; n=26, 49.06% sequenced). Analysis of patient clinical data revealed that 39 (11.57%) of confirmed patients had at least one concomitant sexually transmitted infection other than HIV (including syphilis, gonorrhea, trichomoniasis, hepatitis B), 120 (35.61%) were living with HIV, of which 70 (58.33%) of them reported immunosuppression.

Among the 328 confirmed cases from 2022, we sequenced 50 samples (15%). In 2023, three of the nine confirmed cases were sequenced (33%) **(Figure 1)**. The clinical-epidemiological profile of sequenced cases is presented in **Table 1**. All the 53 MPXV genomes sequenced were assigned by *NextClade* into clade IIb and most of them were assigned to lineage B.1.9 (32/53), followed by B.1.1 (11/53), B.1 (7/53), C.1 (2/53), and B.1.13 (1/53) lineages (**Figure 2A**). Clade and lineage are designated according to the nomenclature proposed by Happi et al. (11).

**Figure 2.**
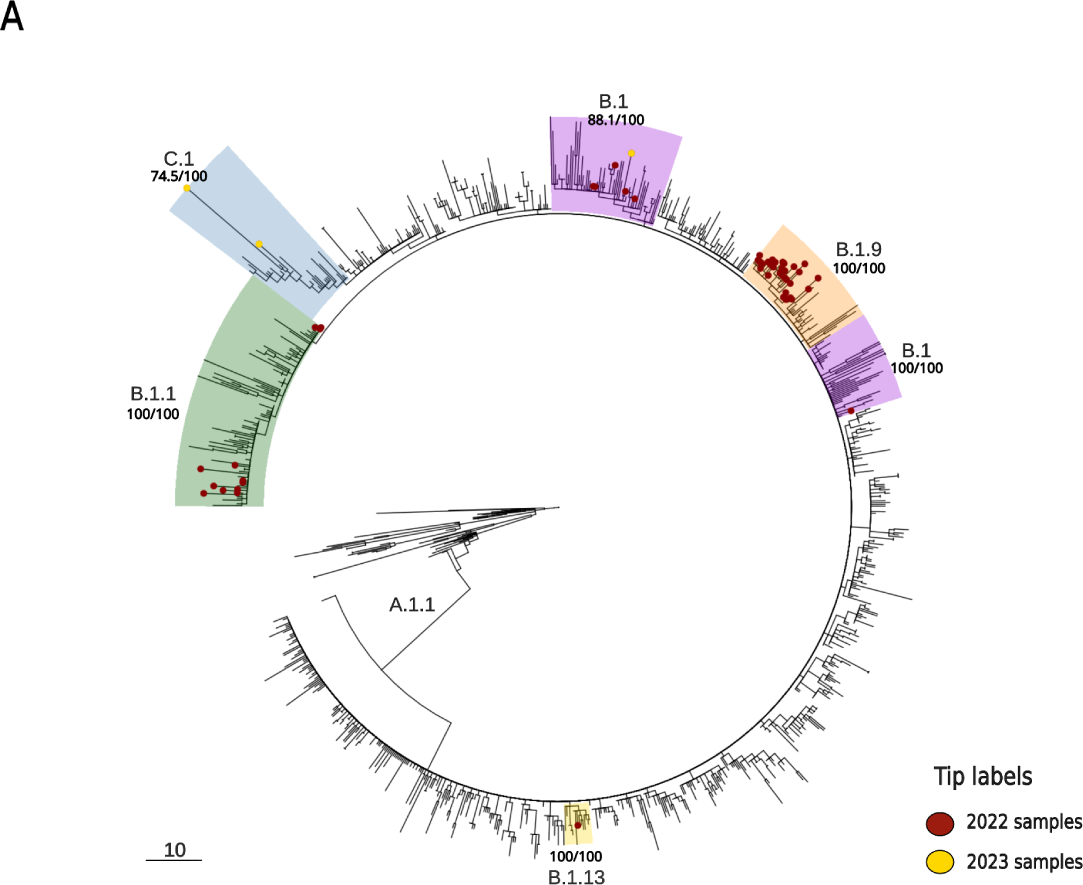
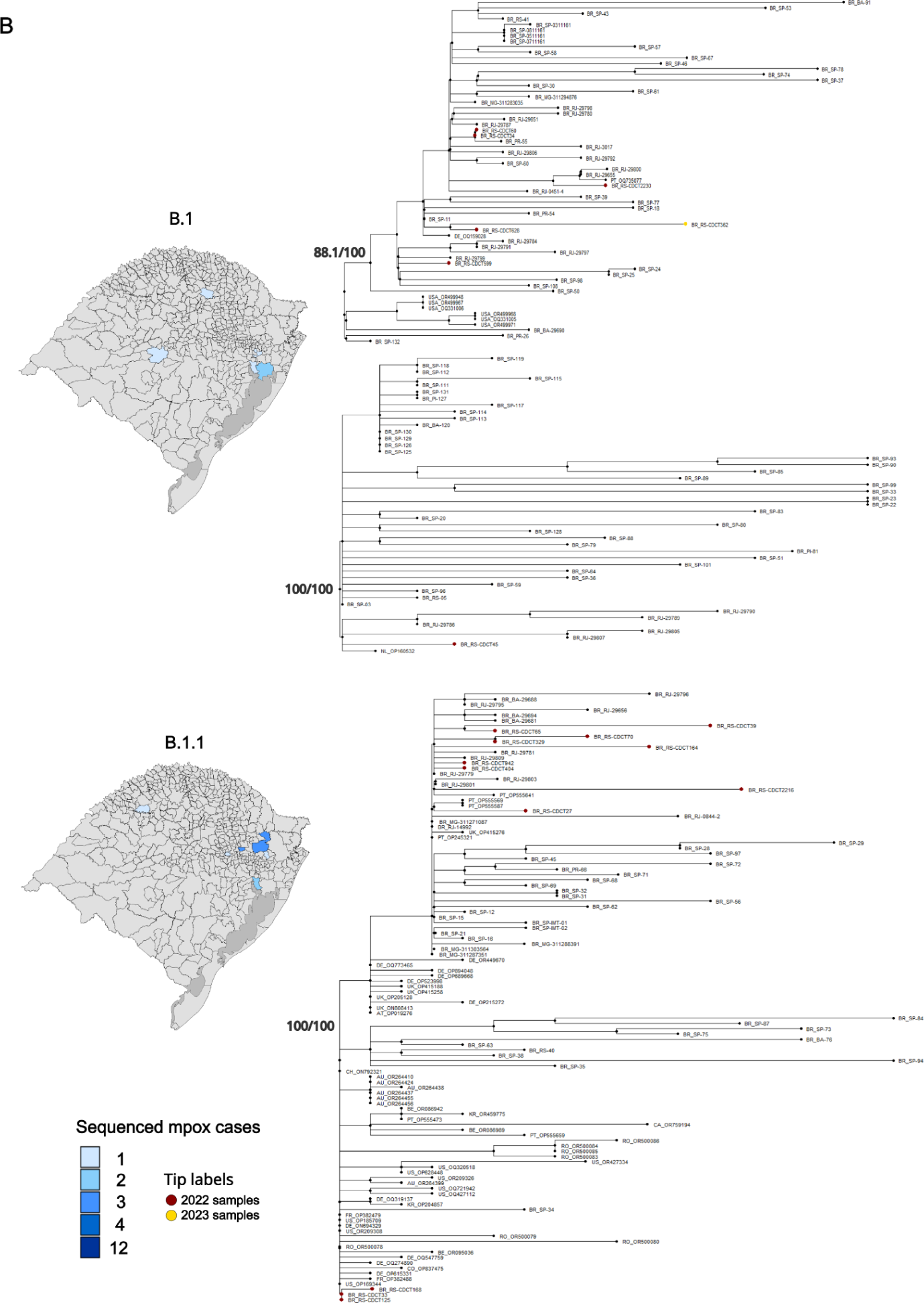
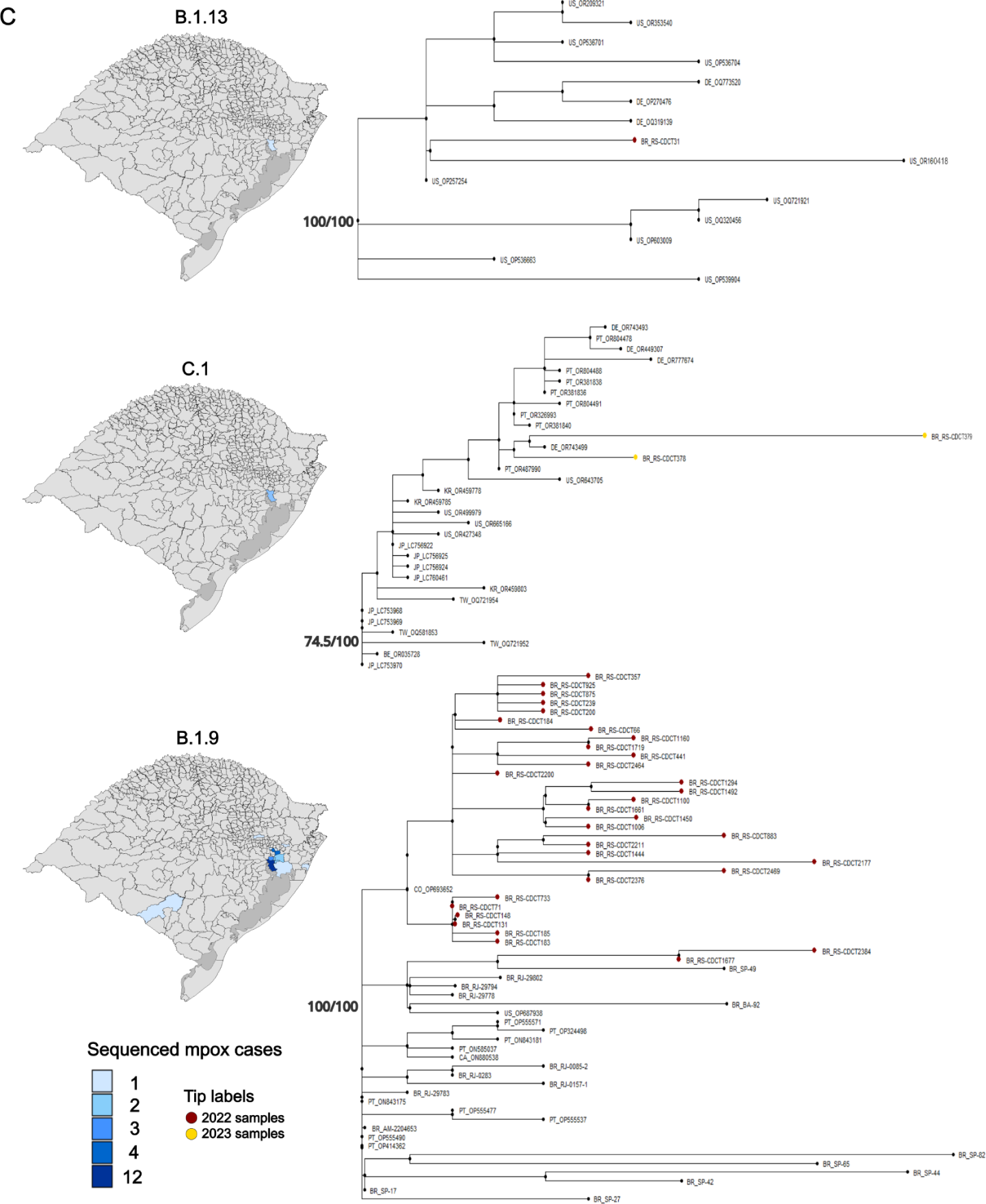
Maximum likelihood tree including the 53 MPXV genomes obtained in this study. The tree incorporates a set of 216 high-quality MPXV sequences sourced from Brazil and a comprehensive dataset of 849 MPXV sequences from diverse global origins (A). The refined presentation in (B) highlights the distinct clades B.1 and B.1, while (C) focuses on the clades B.1.13, C.1, and B.1.9.

Phylogenetic reconstruction supports multiple MPXV international introduction events in RS, from the United States, Portugal, Germany, and Belgium, followed by MPXV community transmission within the state in 2022, the first year of the mpox global spread (**Figure 2**). We identified a genomic cluster that included two sequences from Porto Alegre (capital of the state) residents, with the onset of symptoms one day apart and the same sample collection date, even though no epidemiological link was found among them. Another genomic cluster included samples from an infected patient and a healthcare worker from a possible occupational infection transmitted through fomite exposure to surfaces in the patient’s home (23). Among MPXV genomes from individuals diagnosed in 2023, we identified one case in which the sequence was assigned to the B.1 lineage. Notably, this case clustered in the same phylogenetic clade as an MPXV genome from Rio Grande do Sul sampled in 2022, from a different city, suggesting continued virus transmission in the region (**Figure 2B**). The remaining two genomes from 2023 belonged to the C.1 lineage and were phylogenetically grouped within a clade composed of otherwise Asian and European isolates, including those from Germany and Portugal, suggesting a separate international introduction event (**Figure 2C**).

## Discussion

Consistent with global epidemiological data, MPXV infection incidence was significantly higher in males. While men who have sex with men were the most affected in this outbreak, it is crucial to emphasize that MPXV can infect individuals of all genders and sexual orientations. Therefore, increased surveillance is required to avoid delayed or missed diagnoses when evaluating patients presenting with unusual acute rashes, particularly when accompanied by systemic symptoms (29).

The hospitalization rate of 6.36% observed in our study is slightly lower than those reported by Thornhill et al. (28), who reported a 13% hospitalization rate in a compilation of cases from 16 countries. Cases of mpox in this outbreak were usually characterized by mild symptoms even considering the high proportion of immunocompromised patients. This is a counterintuitive result compared with other reports from the literature where population groups such as children, pregnant women, immunocompromised individuals, individuals living with HIV, or those with concomitant sexually transmitted infections other than HIV experienced more severe symptoms (30,31). It is important to note that clinical manifestations may also be related to the degree and route of exposure to the virus and the patient’s immunological status (32). Moreover, perhaps the immunocompromised patients in Brazil undergo better treatment and follow-up due to the long and successful history of HIV and other sexual infectious diseases treatment in the country. Unfortunately, the available data from the public health system from which the samples are derived does not allow us to further access the long-term treatment information of these patients.

The high proportion of diagnosed cases among men who have sex with men (MSM) in this outbreak could be due to an earlier introduction of MPXV into these interconnected sexual networks. Alternatively, this finding could reflect an ascertainment bias influenced by the pre-existing better relationships between MSM and healthcare providers specializing in sexually transmitted infections (32). However, regardless of geography or specific population groups, close physical contact with infected individuals facilitates mpox transmission, making anyone, irrespective of gender or sexual orientation, susceptible (33). In Brazil, individuals living with HIV demonstrated a 46% higher hospitalization rate compared to those without HIV (34). Similarly, those with concomitant sexually transmitted infections had a 53% increased risk, and the presence of immunosuppression was associated with a 55% higher hospitalization rate. These factors likely contributed to the 16 fatalities recorded in other Brazilian states (24) and emphasize the critical role of individual immunological status in determining the clinical outcome.

Our genomic analysis of the MPXV in Rio Grande do Sul, Brazil during the 2022 outbreak and its sustained circulation in 2023 revealed several important insights into MPXV introduction and transmission dynamics. The data provide valuable evidence about the initial virus introduction and the ongoing circulation of the virus. Our findings regarding clustered sequences originating from diverse global locations, including the United States, Portugal, Germany, and Belgium demonstrated the multiple independent introductions into Rio Grande do Sul during the early phases of the outbreak. The initial case identified in Rio Grande do Sul was recorded in May 2022, coinciding with the onset of the first outbreak cases reported in Europe and North America (6,7,34). This chronologically proximate occurrence emphasizes the rapid MPXV transmission across different continents, facilitated by the interconnectedness of global populations and the continuous movement of infectious agents across borders. These findings corroborate the importance of international collaboration in monitoring, reporting, and responding to emerging infectious diseases to prevent and mitigate their impact on a global scale (35).

The dominance of lineage B.1.9 within the sequenced samples, alongside indications of local community transmission, strongly indicates the successful establishment of the virus in the state. First reported in Portugal, this lineage exhibited a notable prevalence in the European country in the first year of the global outbreak, when 93% of all B.1.9 sequences available were from Portugal (36). In the following months after first detection in Portugal, this lineage was introduced and disseminated across Brazil, where now most of the genomes sequenced belong to this lineage, as 51 out of 103 sequences assigned to lineage B.1.9 currently available in *GISAID* were sampled in Brazil. Regarding the identification of two distinct but geographically close genomic clusters within our dataset, while no established epidemiological link was found for the first cluster, the second cluster involved a patient and a healthcare worker indicating the possibility of nosocomial transmission through environmental fomites, which underscores the importance of infection control measures and personal protective equipment (PPE) use in healthcare settings (23).

The MPXV assigned to lineage B.1 from a sample collected in September 2023, clustering with a sequence from Rio Grande do Sul collected in 2022, suggests the ongoing local transmission of MPXV in the region (37). On the other hand, the two C.1 lineage sequences identified in mpox cases diagnosed in August and November 2023, were grouped within a monophyletic lineage dominated by sequences sampled in Asia and Europa sequences indicating a new introduction event potentially occurring later in the outbreak. These findings underscore the recurrence of introductions and emphasize the continuous need for genomic surveillance to effectively track circulating lineages to inform public health interventions (14). Future efforts should prioritize increased sequencing for a more comprehensive understanding of viral diversity and transmission dynamics. Viral genomics from well-sampled cases alongside epidemiological investigations provide invaluable insights into specific transmission routes and inform targeted control measures. Learning from mpox, and enhancing global health systems surveillance and cooperation can improve preparedness and reduce the impact of future health threats. This proactive approach, inspired by the successful strategies employed in the surveillance of diseases like COVID-19, allows for the early detection of variants and the timely implementation of targeted public health interventions (38, 39).

The long-term maintenance of the MPXV virus, as indicated in this study, suggests the possibility that the virus has been circulating within Rio Grande do Sul for an extended period, potentially establishing itself and significantly increasing the chance of evolving into an endemic pathogen. This scenario could be due to factors like undetected chains of transmission or a significant number of asymptomatic cases. Understanding and addressing these factors are crucial for effective management and control of the virus transmission in the community. In conclusion, our results provide valuable insights into the multiple introductions, community transmission, and ongoing long-term circulation of mpox in Rio Grande do Sul. Our findings stress the importance of genomic surveillance to guide infection control measures such as the identification of occupational transmission chains, mode of transmission, and hidden transmission chains to mitigate the spread of this emerging virus.

Other mpox control measures include improving surveillance systems for early case detection and reporting, ensuring rapid and accurate laboratory testing for confirmation, and actively monitoring close contacts of confirmed cases. Additionally, public awareness campaigns are crucial to informing the population about mpox symptoms, transmission pathways, and effective prevention strategies. Further research, incorporating increased sequencing and detailed epidemiological investigations, is also crucial to thoroughly understand the international and regional MPXV transmission dynamics and to inform effective public health responses.

## Acknowledgments

This work was supported by Fundação de Amparo à Pesquisa do Estado do Rio Grande do Sul (FAPERGS/MS/CNPq 08/2020–PPSUS, Grant process 21/2551-0000059-7, FAPERGS/FIOCRUZ 13/2022 – REDE SAÚDE-RS, grant process 23/2551-0000510-7 and FAPERGS 14/2022 - ARD/ARC, grant process 23/2551-0000852-1). Conselho Nacional de Desenvolvimento Científico e Tecnológico (CNPQ) and Fundação de Amparo à Pesquisa do Estado do Rio Grande do Sul has provided a fellowship to R.S.S (FAPERGS/CNPq 07/2022) and a productivity grant PQ1D to GLW (307209/2023-7). ABGV holds a Research Fellowship (PQ2) from CNPq (Grant process 304476/2022-6).

## Credit authorship contribution statement

Fernanda Godinho: Conceptualization, Methodology, Investigation, Writing - Review & Editing. Mayara M. Oliveira: Conceptualization, Methodology, Investigation, Writing - Review & Editing. Regina B. Barcellos: Methodology, Investigation. Amanda P. Ruivo: Methodology, Investigation. Viviane H. de Melo: Methodology, Investigation. Thales Bermann: Investigation, Writing – review & editing. Franciellen M. Santos: Methodology, Investigation. Milena Bauerman: Methodology, Investigation. Taina M. Selayaran: Methodology, Investigation, Software. Taina S. Soares: Methodology, Investigation. Patrícia Sesterheim: Methodology, Investigation. Ludmila F. Baethgen: Investigation, Writing – review & editing. Fernanda M. da Rocha: Data curation, Methodology. Renata P. Mondini: Data curation, Methodology. Karine M. Amaral: Data curation, Methodology. Fernanda C. L. Delela: Data curation, Methodology. Sabrina Vizeu: Data curation, Methodology. Tatiana S. Gregianini: Data curation, Methodology. Ana B.G. Veiga: Funding acquisition, Writing – review & editing. Gabriel L. Wallau: Supervision, Conceptualization, Investigation, Writing – review & editing. Richard S. Salvato: Supervision, Conceptualization, Investigation, Writing – review & editing.

## Ethics approval

This project was approved by the Research Ethics Committee (CEP) at Escola de Saúde Pública (SES-RS). Process number: CAAE: 67181123.1.0000.5312.

## Data availability

Consensus sequences are available in the GISAID database under accession codes: EPI_ISL_14465517, EPI_ISL_14676265, EPI_ISL_15165602 - EPI_ISL_15165618, EPI_ISL_17406093 - EPI_ISL_17406098, EPI_ISL_17406100 - EPI_ISL_17406124, EPI_ISL_18971016 - EPI_ISL_18971018.

## Supplementary information

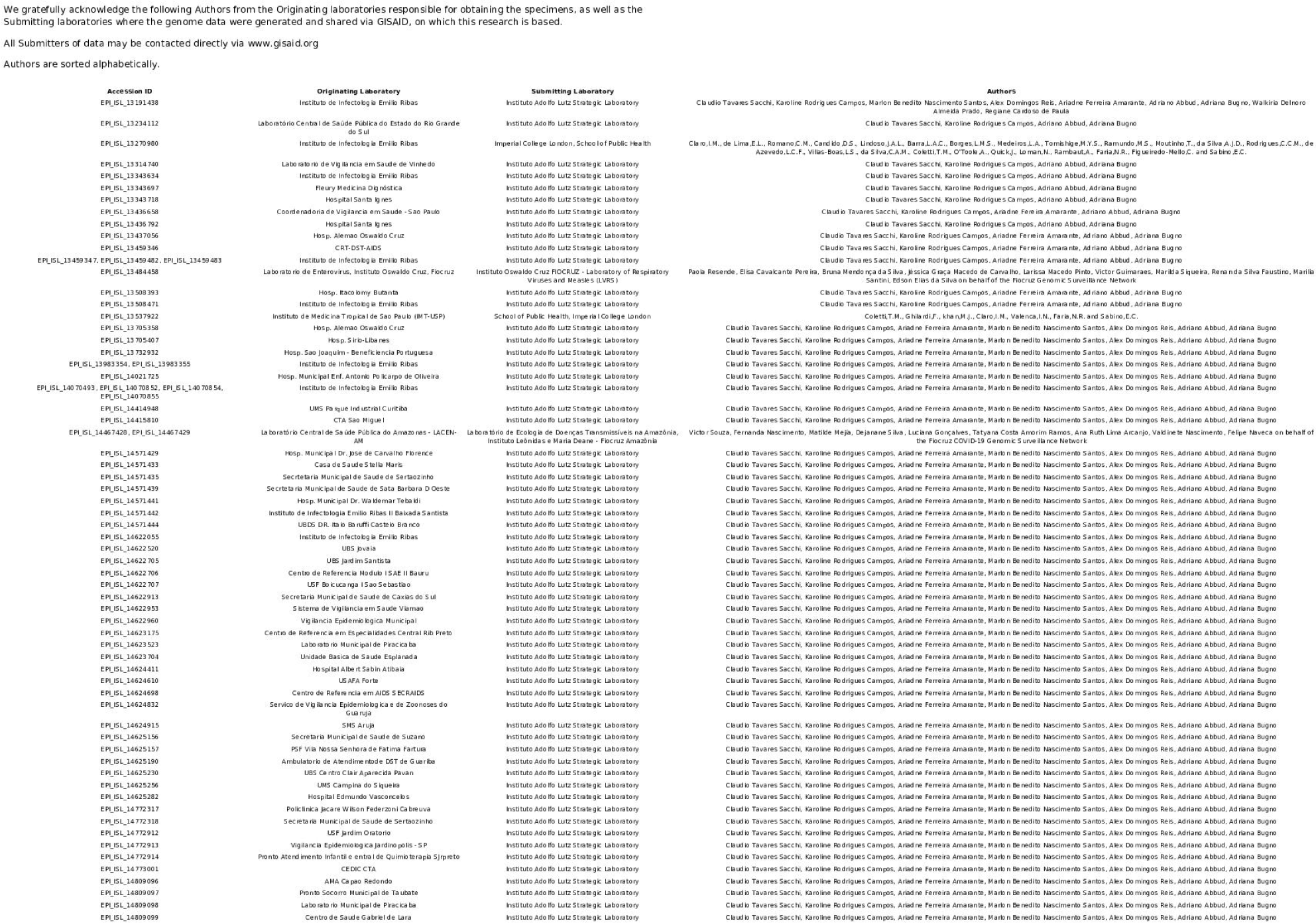
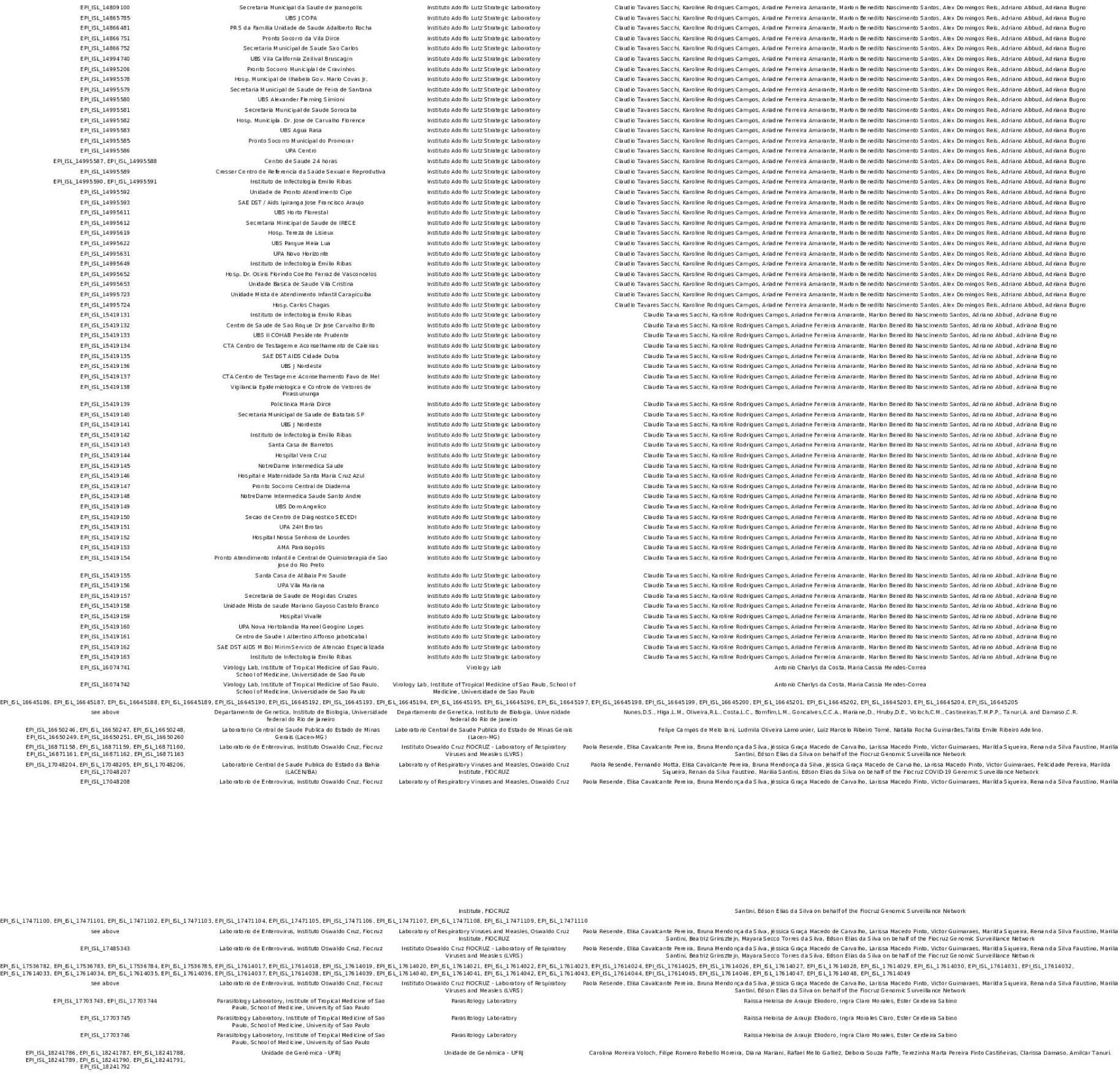

